# Quantifying the causal impact of biological risk factors on healthcare costs

**DOI:** 10.1101/2022.11.19.22282356

**Authors:** Jiwoo Lee, Sakari Jukarainen, Padraig Dixon, Neil M Davies, George Davey Smith, Pradeep Natarajan, Andrea Ganna

## Abstract

**Background:** A critical step in evaluating healthcare interventions is to understand their impact on healthcare costs. However, there is a limited understanding of the causal impact that biomarkers and risk factors for disease have on healthcare-related costs. Previous studies based on observational data have major limitations including residual confounding and reverse causation. Here, we used a genetically-informed design, Mendelian Randomization (MR), to infer the causal impact of 15 routinely measured and clinically relevant risk factors on annual total healthcare costs.

**Methods:** We considered 373,160 participants from the FinnGen Study, which were linked to detailed healthcare costs covering inpatient, outpatient, and medication costs. Several MR approaches were used to assess the causal effects of 15 risk factors (e.g., waist circumference (WC), HDL cholesterol, vitamin D), with strong genetic bases on annual total healthcare costs, as well as stratified by service type, age, and sex. We further assessed the generalizability and robustness of our results by accounting for selection bias and by leveraging additional data from 323,774 individuals from the United Kingdom and Netherlands.

**Results:** Robust causal effects were observed for waist circumference (WC), adult body mass index, and systolic blood pressure, in which a one standard deviation increase in the risk factors corresponded to 22.78% [95% CI: 18.75, 26.95], 13.64% [10.26, 17.12], and 13.08% [8.84, 17.48] increased annual total healthcare costs, respectively. The relative effect of WC on annual total healthcare costs was consistent across age and sex and was not attenuated when accounting for increased risk of five major diseases: back pain, chronic ischemic heart disease, type 2 diabetes, chronic obstructive pulmonary disease, and stroke. A lack of causal effects was observed for some clinically relevant biomarkers, such as albumin, C-reactive protein, and vitamin D.

**Conclusion:** Our results indicated that increased WC is a major contributor to annual total healthcare costs and more attention should be given to WC screening, surveillance, and mitigation. On the contrary, several biomarkers relevant in clinical settings did not have a direct impact on annual total healthcare costs.

## Introduction

Healthcare costs continue to rise worldwide, and in 2018, global healthcare spending reached $8.3 trillion, or 10% of the global gross domestic product.^1^ While healthcare costs continue to rise, morbidity is rising, so better understanding of healthcare costs and cost efficiency is critical.^1^ Accurate measurement of healthcare costs caused by different risk factors and health outcomes is important to prioritize public health promotion and prevention programs.^2^ Moreover, healthcare costs can act as a proxy of disease burden when investigating the effects of risk factors. Thus, epidemiology, public health, and policy stakeholders are very interested in the analysis of healthcare costs.^3^

Several studies have quantified the healthcare costs associated with different risk factors.^4 5^ For example, Bolnick et al.^4^ calculated the correlation between United States healthcare spending and 84 modifiable risk factors from the Global Burden of Disease study, and Goetzel et al.^5^ calculated the correlation between healthcare costs and 10 modifiable risk factors including blood glucose, obesity, stress, depression, and physical inactivity. However, there are several limitations with such studies. First, associations between risk factors and healthcare burden are based on observational data and suffer from challenges such as confounding and reverse causation. Second, most studies do not estimate the direct association between risk factors and healthcare costs, but first estimate the impact of risk factors on different diseases and subsequently link each disease to estimated healthcare costs.^4 6 7^ Thus, the impact of risk factors on healthcare costs that are not directly captured by diseases (e.g., medications) were not considered. Third, while modifiable risk factors such as smoking and alcohol consumption have been studied,^8^ little is known about the impact on healthcare costs of commonly measured biomarkers, which are generally the direct targets of pharmacological interventions.

An alternative source of evidence to assess the effects of diseases and biomarkers on healthcare costs is Mendelian Randomization (MR), which addresses some of the previous limitations. MR is a method that uses genetic variants as instrumental variables to estimate causal relationships between exposures and outcomes and can address the issues of confounding and reverse causation.^9^ MR is particularly powerful for estimating the effects of biological risk factors with a strong genetic bases, such as clinical biomarkers and biometrics, including body mass index and blood pressure.

Previous studies have used MR to identify the causal effects of adiposity,^10^ body mass index,^11,12^ and common health conditions.^13^ However, these studies were either based in the UK Biobank (e.g., limited to relatively healthy individuals between 40 and 69 years old) or did not have complete coverage of healthcare costs associated with medication and primary care costs. No studies to date have used MR to comprehensively link a diverse set of biological risk factors to healthcare costs. In this study, we used a large prospective study from Finland, the FinnGen Study, with genetic information available for 373,160 individuals linked to several national health registries covering primary, secondary, and medication costs. Because of the high-quality, long follow-up, and detailed healthcare costs available in these registries, we were able to obtain an accurate and comprehensive estimate of annual healthcare expenditure. We further assessed the generalizability and robustness of our findings by accounting for selection bias and by leveraging additional healthcare cost data from 323,774 individuals from the United Kingdom and Netherlands.

In this study, we aimed to (1) evaluate the causal impact of 15 risk factors, with strong genetic bases, on annual total healthcare costs, (2) identify whether the effects vary by service type, age, and sex, and (3) quantify the mediating of effects of major diseases.

## Methods

### Study cohort

This study utilized data from the FinnGen Study, which is an ongoing prospective cohort study aiming to recruit 520,000 individuals by combining population-based legacy cohorts, disease-based cohorts, and volunteers recruited by biobanks.^14^ The average age at baseline (i.e., date of DNA sample collection) is 54 years old and 56% of the study cohort is female. Participants are linked to national health registries that provide rich longitudinal information. Such registries include the Register of Primary Health Care Visits (AvoHILMO) which captures outpatient visits, the Care Register for Health Care (HILMO) which captures hospital visits, and the Medication Reimbursement Register (Kela). Individual-level genotypes and register data from FinnGen participants can be accessed by approved researchers via the Fin-genious portal (https://site.fingenious.fi/en/) hosted by the Finnish Biobank Cooperative FinBB (https://finbb.fi/en/). Data release to FinBB is timed to the bi-annual public release of FG summary results which occurs twelve months after FG consortium members can start working with the data.

Given that the study participants in FinnGen may differ from the entire Finnish population due to its hospital-based recruitment (e.g., individuals in FinnGen are typically sicker and have higher disease prevalence), we adjusted the study cohort in FinnGen to the entire Finnish population using inverse probability weights in a subsequent sensitivity analysis. We used the calibration weighting method, which uses the marginal proportions of variables to adjust the sample weights to satisfy the population margins. We used the following five health and sociodemographic characteristics: age, gender, education, occupation, and region of birth.

### Estimation of healthcare costs

(1) AvoHILMO and (2) HILMO are registries maintained by the Finnish Institute for Health and Welfare (THL) for (1) primary outpatient and (2) secondary and tertiary inpatient and outpatient hospital visits, respectively. The Finnish Institute for Health and Welfare publishes average unit cost estimates for different types of healthcare services (e.g., outpatient visits, inpatient episodes). The Social Insurance Institution (SII, also known as Kela), the Finnish government agency in charge of national social security programs, maintains a registry of all reimbursed prescription medication purchases in Finland. The AvoHILMO registry was started in 2011, the HILMO registry in 1998, and the medication purchases registry in 1998. All AvoHILMO, HILMO, and medication costs capture total costs regardless of the payer. We did not capture costs related to non-reimbursed medications and home care. Going forward, we referred to any AvoHILMO costs as “primary care costs”, HILMO costs as “secondary care costs”, and Kela costs as “medication costs”.

We used the unit cost estimates published by the Finnish Institute for Health and Welfare to obtain costs associated to each medical encounter.^15,16^ Primary care costs were linked to each medical encounter by profession (e.g., physician, nurse), service type (e.g., primary healthcare, mental health), and contact type (e.g., visit, phone call). Secondary care costs were linked based on service (e.g., emergency room visit, outpatient visit, inpatient visit), specialty (e.g., cardiology, neurology), and hospital (e.g., university, central, other) types. Medication costs were linked using the Nordic Article Number (VNR), which is an identifier that exactly captures the type of medicinal product (e.g., manufacturer, dosage) purchased. We used the yearly average costs for each VNR code across Finnish pharmacies to link the costs. Primary care costs prior to 2011 were excluded, and secondary care and medications costs prior to 1998 were excluded to reflect the start dates of each registry. Individuals with secondary care or medication records, but without primary care records, were assumed to be individuals using private primary healthcare services. There were 294 (0.08%) such individuals in FinnGen, and they were assigned the median primary care cost of €71.62. For all cost categories, we examine costs in 2017 euro values such that the same service contributes similarly to costs whether it occurred for example in 2010 or 2017. Other missing values were assigned zero values (i.e., individuals with primary and secondary care records, but without medication records, were assigned a zero value for medication costs). To adjust for fluctuating healthcare costs by different years, each unique set of identifiers was assigned to the standardized healthcare costs in 2017.

We estimated the annual total healthcare costs, primary care, secondary care, and medication costs adjusted by the total follow-up time that individuals were observed in each registry. The start of follow-up was defined as 2011 for AvoHILMO and 1998 for HILMO and Kela. The end of follow-up (EOF) was defined as date of death, date of emigration, or the end-of-registry date (October 11, 2021). The annual total healthcare costs for each individual are estimated as:

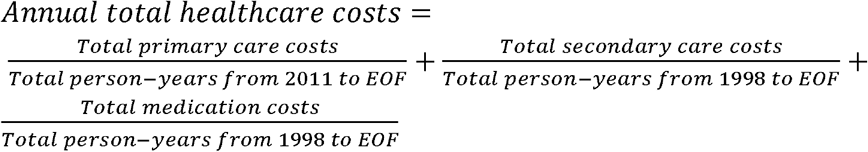

As healthcare costs were highly right-skewed in this sample, a log(X + 1) transformation was implemented before modeling. Annual total healthcare costs were main outcome studied. In sensitivity analyses, we examined healthcare costs stratified by: (1) service type (e.g., primary care, secondary care, and medication costs), (2) sex, and (3) age (individuals under 30 years old, individuals between 30 and 60 years old, and individual over 60 years old).

### Mendelian Randomization

We performed MR, which is a method that uses genetic variants as instrumental variables to estimate the effect of specific exposures on healthcare costs.^9^ The exposures included 15 biological risk factors based on the following criteria: (1) has strong genetic instruments (e.g., F-statistic > 50) and (2) of clinical interest and relevance (e.g., can be measured through available laboratory tests). We used summary statistics from the GWAS of healthcare costs conducted in FinnGen for the outcomes and non-overlapping summary statistics from the MRC IEU OpenGWAS Database for the exposures (**Supplementary Table 1**).^17^ Some summary statistics were back-transformed from standardized to raw units on the original scale (e.g., adult body mass index, HDL cholesterol, LDL cholesterol, triglycerides, systolic blood pressure, and waist circumference).

The primary outcome was log-transformed annual total healthcare costs, and our secondary outcomes included (1) log-transformed primary care, secondary care, and medication costs, (2) log-transformed annual total healthcare costs for females and males, and (3) log-transformed annual total healthcare costs for individuals under 30 years old, individuals between 30 and 60 years old, and individuals over 60 years old. We performed genome-wide association studies (GWAS) of healthcare costs to identify genetic variants associated with healthcare costs using REGENIE, which is a method for fitting a whole-genome regression model.^18^ Briefly, REGENIE uses a two-step process that fits a whole-genome regression model and performs single-variant association testing. We used the default model with the following covariates: birth year, birth year squared, sex, 10 principal components, and batch covariates.

To evaluate the causal effect of the 15 risk factors on healthcare costs, we performed two-sample MR, which utilizes summary statistics from GWAS of exposures and outcomes in non-overlapping cohorts.^19^ MR relies on several assumptions: (1) genetic instruments must be robustly associated with the exposure, (2) there must be no confounders of the genetic instruments-cost associations, and (3) genetic instruments must not influence costs except through the exposure of interest.^9^ We performed two-sample MR using the TwoSampleMR package in R.^17,20^

We performed LD clumping with a window of 10000 kilobases and an R^2^ cutoff of 0.002 and utilized the MR Egger, weighted median, inverse variance weighted, simple mode, and weighted mode methods. The inverse variance weighted method estimates the causal effect based on a ratio of association estimates from a univariable regression of the outcome on the genetic variant and the exposure on the genetic variant, averaging each ratio estimate with inverse variance weights. The MR Egger method uses a similar method, with the exclusion of an intercept term. The simple mode, weighted mode, and weighted median methods rely on similar approaches, with different weights. Several methods were used in combination due to differing advantages and disadvantages. For example, the MR Egger method, is more robust to pleiotropy (e.g., one variant affecting multiple phenotypes), yet suffers from lack of power, while the inverse variance weighted method retains more statistical power.

### Multivariate Mendelian Randomization

Multivariate MR (MVMR) uses genetic variants for two or more exposures to simultaneously estimate the causal effect of each exposure on the outcome, controlling for the effect of the other included exposures. MVMR can therefore use genetic variants for several risk factors to estimate independent and direct effects of these risk factors, as well as estimating mediation.^21^ MVMR requires the same assumptions as univariate MR, but the genetic instruments must be associated with the set of exposures rather than the single exposures, but it is not necessary for each genetic instrument to be associated with every exposure.^21^

We performed MVMR to identify mediators of the exposures on healthcare costs, in which the mediators were the top five noncommunicable diseases from the Global Burden of Disease:^22^ back pain, chronic ischemic heart disease, type 2 diabetes, chronic obstructive pulmonary disease, and stroke. Summary statistics for mediators were obtained from the UK Biobank (**Supplementary Table 1**). For example, we estimated the simultaneous effects of chronic ischemic heart disease and waist circumference on healthcare costs by including two exposures in our model, which were identified using genetic variants associated with each exposure. This allowed us to obtain the direct effect of waist circumference on healthcare costs by adjusting out the indirect effect of chronic ischemic heart disease on healthcare costs.

### Replication analyses in the United Kingdom and Netherlands

We performed validation analyses in the UK Biobank ^23^ (N=307,048) and Netherlands Twin Register ^24^ (N=16,726) to evaluate the generalizability and robustness of our results to different healthcare systems in different countries. For the Netherlands Twin Register, we used published GWAS summary statistics ^24^. For UK Biobank, we expand upon published analyses for BMI ^23^. We repeated our analyses to estimate the annual monetary impact per capita associated with each risk factor. For the UK Biobank, we converted pounds to euros using the average exchange rate in 2021 of 1 EUR = 0.8403 GBP. We also calculated the genetic correlation between the three sets of summary statistics to evaluate the consistency of associations across different healthcare systems using LDSC, a tool to estimate genetic correlation and heritability.^25^ We also constructed polygenic scores of healthcare costs from the UK Biobank and Netherlands Twin Register and estimated their associations with healthcare costs in FinnGen. We first used PRS-CS ^26^ to calculate weights of association and PLINK2 ^27^ to calculate scores. Briefly, PRS-CS is a polygenic prediction method that uses Bayesian regression and infers posterior SNP effect sizes under continuous shrinkage priors using only GWAS summary statistics and an external linkage disequilibrium reference panel.^26^ The 1000 Genomes EUR reference panel was used to output weights using standard PRS-CS parameters. Only HapMap3 variants were included. PLINK2 was used to calculate the polygenic scores, which were then standardized across the entire FinnGen Study cohort with a mean of 0 and a standard deviation of 1.

## Results

In this study (**Figure 1**), we estimated the causal impact of 15 risk factors with strong genetic bases (**Supplementary Table 1**) on annual total healthcare costs.

**Figure 1.**
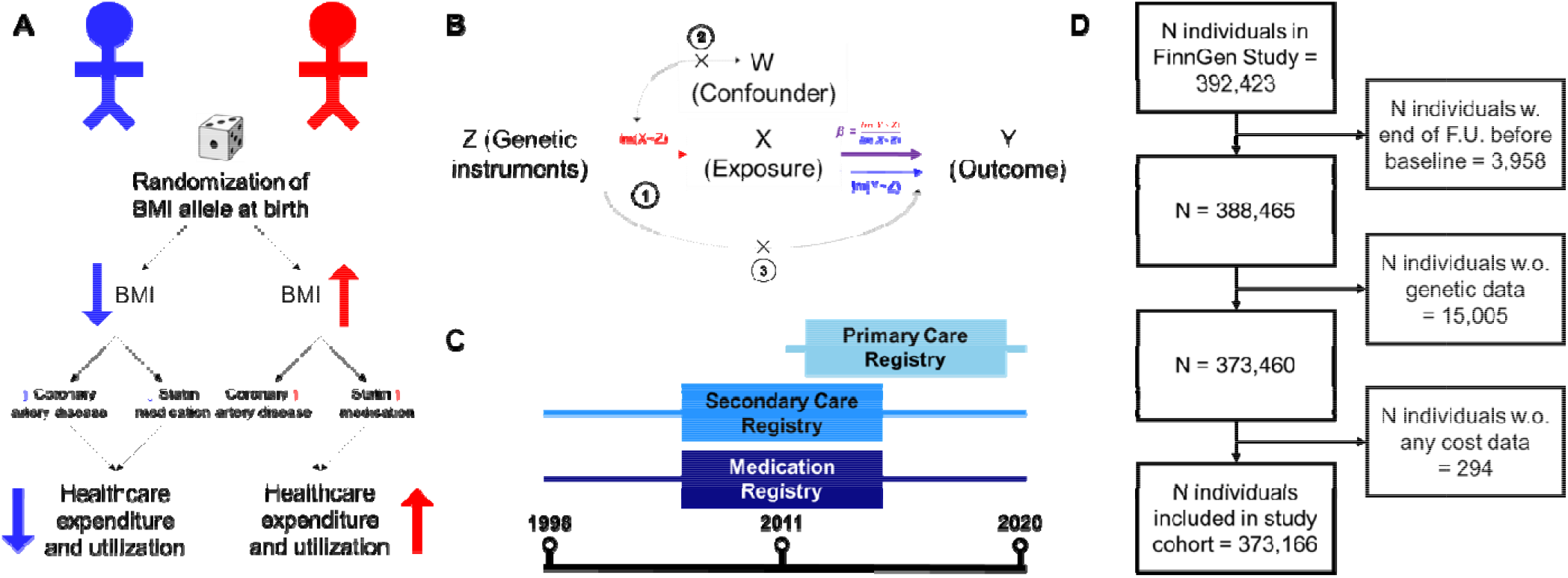
Graphical abstract. (A): Example of how genetic variants associated with BMI and randomly assigned at birth can be used to infer the causal impact of BMI on healthcare costs (e.g., by modifying risk for cardiovascular disease and statin medication). (B): Assumptions underlying MR. 1: Genetic instruments must be robustly associated with the exposure (risk factor), 2: there must be no confounders of the genetic instruments-outcome association, and 3: Genetic instruments must not influence the outcome except through the exposure. (C): National healthcare registries link with the FinnGen Study to estimate annual total healthcare costs. (D) STROBE flow diagram for study cohort, in which 373,160 individuals were included.

### Distribution of healthcare costs

We included 373,160 FinnGen participants (data freeze 8) followed-up to a maximum of 22 years. The average age at baseline (i.e., date of DNA sample collection) was 54 years old and 56% of the study cohort was female. The mean and median annual total healthcare cost was €2,706 and €1,313, respectively (**Figure 2**). Primary care (mean = €169, median = €109) and medication (mean = €518, median = €202) costs were lower than secondary care (mean = €2019, median = €852) costs. Mean (females = €2244, males = €3303) and median (females = €1245, males = €1433) costs were similar in male and females but males (SD = €15545) had greater variability than females (SD = €4445). Individuals over the age of 60 (mean = €3406, median = €1800) had greater healthcare costs than individuals between the age of 30 and 60 (mean = €1851, median = €891) and individuals under the age of 30 (mean = €1484, median = €621).

**Figure 2.**
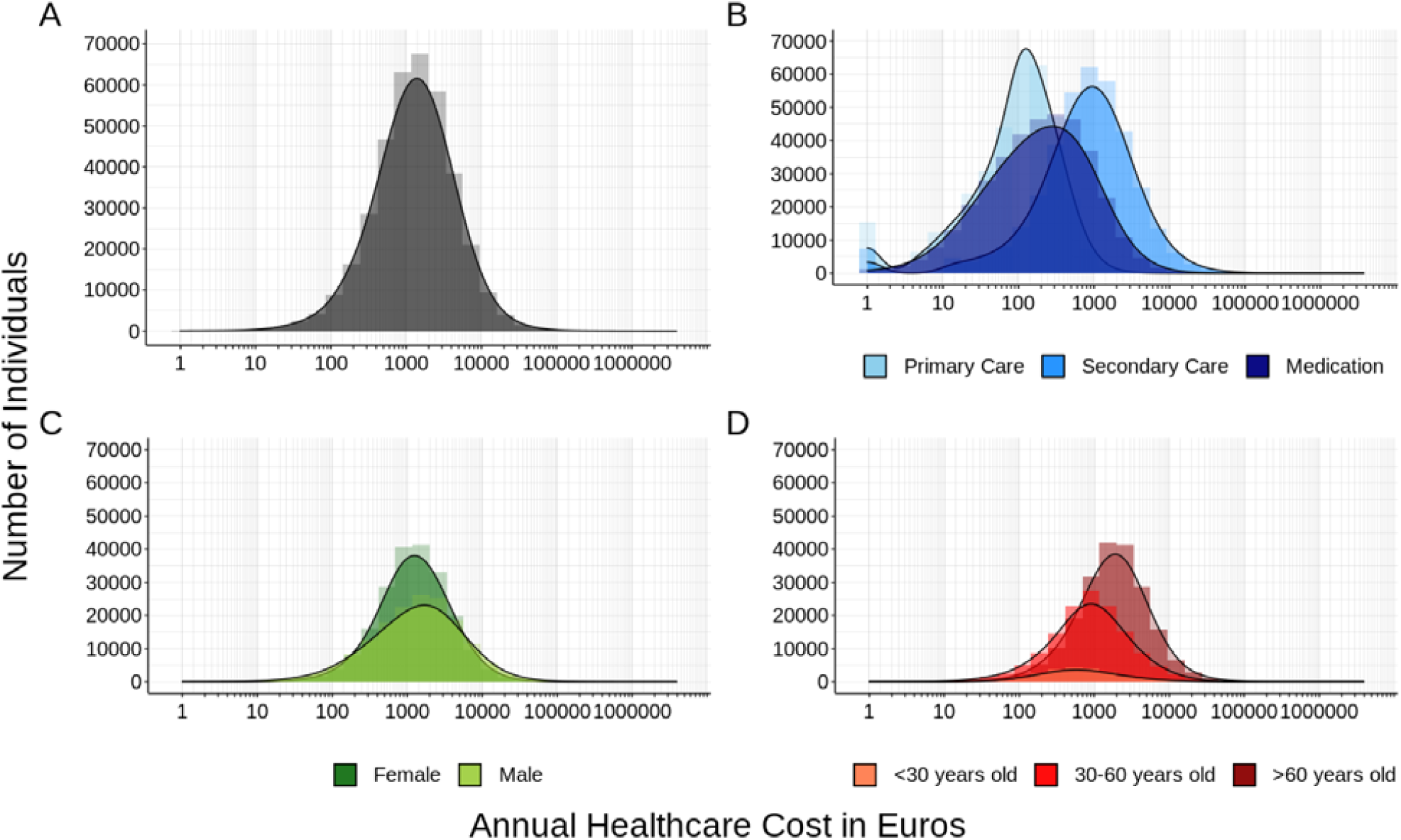
Distribution of healthcare costs in 373,160 FinnGen participants. (A) Annual total healthcare cost in euros. (B) Annual healthcare costs in euros for primary care, secondary care, and medication costs. (C) Annual total healthcare costs in euros for females and males. (D) Annual total healthcare costs for individuals under 30 years old, between 30 and 60 years old, and over 60 years old. X-axis is on a log10-transformed scale.

### Causal impact of risk factors on total healthcare costs

We estimated the causal impact of risk factors on healthcare costs using MR. All risk factors had strong genetic instruments (e.g., F-statistic > 50) obtained from genome-wide association studies of at least 173,082 individuals. We detected significant effects of six risk factors on costs (i.e., waist circumference, adult body mass index, systolic blood pressure, triglycerides, cystatin C, and HDL cholesterol) at the Bonferroni-corrected significance level (P < 3.33×10^−3^) (**Figure 3**). We performed sensitivity analyses using five different robust MR approaches (**Supplementary Table 2**) and identified three risk factors that consistently affected total annual healthcare costs across at least three of the sensitivity analyses: waist circumference (WC), adult body mass index (BMI), and systolic blood pressure (SBP). One standard deviation (SD) increase in WC increased the annual total healthcare costs by 22.78% (95% CI: [18.75, 26.95], P = 1.90×10^−33^); one SD increase in adult BMI increased the annual total healthcare costs by 13.64% (95% CI: [10.26, 17.12], P = 1.06×10^−16^); and one SD increase in SBP increased the annual total healthcare costs by 13.08% (95% CI: [8.84, 17.48], P = 2.80×10^−10^). Using MR methods robust to pleiotropy, we found similar effects for WC (17.19% - 22.78%), adult BMI (8.21% - 13.64%), and SBP (13.08% - 32.36) (**Supplementary Table 2**).

**Figure 3.**
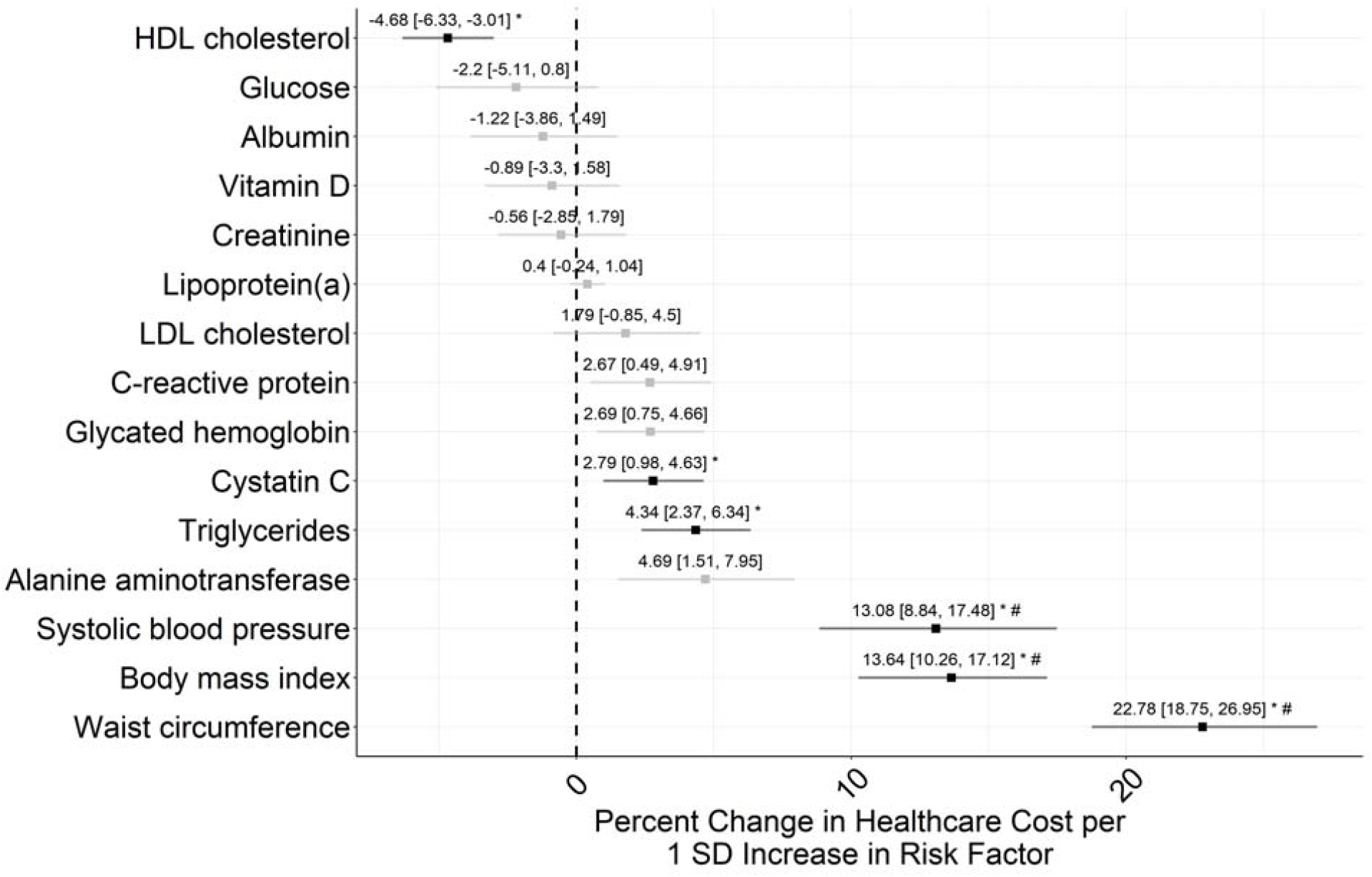
Mendelian Randomization on 15 biological risk factors on annual total healthcare costs for 373,160 FinnGen participants using the two-sample, inverse variance weighted approach. Bars indicate 95% confidence interval. Black bars and the * symbol indicate biological risk factors that are statistically significant at the Bonferroni-corrected significance level (P < 3.33×10^−3^). The # symbol indicates biological risk factors that were significant across at least three of the MR approaches used in sensitivity analyses. SD is standard deviation.

Several biomarkers did not have a significant (e.g., Bonferroni-corrected significance level of P < 3.33×10^−3^) impact on annual total healthcare costs (e.g., alanine aminotransferase, P = 4.58×10^−2^; glycated hemoglobin, P = 6.44×10^−3^; C-reactive protein, P = 1.64×10^−2^; LDL cholesterol, P = 1.86×10^−1^; lipoprotein(a), P = 2.20×10^−1^; creatinine, P = 6.39×10^−1^; vitamin D, P = 4.75×10^−1^; albumin, P = 3.73×10^−1^; glucose, P = 1.50×10^−1^), indicating that genetically-increased levels of these biomarkers do not result in a significant downstream impact on healthcare costs. LDL cholesterol (1.79%, 95% CI: [-0.85, 4.50], P = 1.86×10^−1^) had a null effect on healthcare costs, despite the strong genetic instruments for LDL cholesterol. We performed sensitivity analyses using genetic instruments for triglycerides, HDL cholesterol, and LDL cholesterol that were adjusted for statin usage and observed similar results (**Supplementary Table 2**)

### Impact of risk factors on total healthcare costs

To quantify the amount of annual total healthcare costs associated with WC, adult BMI, and SBP in absolute euros (instead of percent changes), we assumed a median annual total healthcare cost of €1312.53 (**Table 1**). One SD increase in WC, adult BMI, and SBP resulted in increases of €298.99, €179.03 and €171.68 of annual total healthcare costs, respectively. Using clinically interpretable units, we estimated €202.13 annual increase per additional 10 cm of WC; €178.51 per 5 kg/m^2 of adult BMI; and €84.00 increase per 10 mmHg of SBP.

**Table 1.** Monetary impact of three main biological risk factors for 343,160 FinnGen participants as estimated from Mendelian Randomization. SD is standard deviation.

| Exposure | Percent change in cost per unit change in exposure | Confidence Interval | Estimated absolute change in euros |
| --- | --- | --- | --- |
| Waist circumference | 22.78 per 1 SD | [18.75, 26.95] | €298.99 per 1 SD |
|  | 15.40 per 10 cm | [12.90, 17.90] | €202.13 per 10 cm |
| Body mass index | 13.64 per 1 SD | [10.26, 17.12] | **€179.03 per 1 SD** |
|  | 2.72 per 5 kg/m<sup>2</sup> | [2.07, 3.37] | **€178.51 per 5 kg/m<sup>2</sup>** |
| Systolic blood pressure | 13.08 per 1 SD | [8.84, 17.48] | **€171.68 per 1 SD** |
|  | 0.64 per 10 mmHg | [0.44, 0.84] | **€84.00 per 10 mmHg** |

### Impact of risk factors on total healthcare costs by service type, sex, and age

We quantified the impact of six risk factors with significant effects on annual total healthcare costs by repeating the analyses by each service type (i.e., primary care, secondary care, medication), sex, and age (**Figure 4**). SBP (medication vs. secondary care costs, P = 5.75×10^−9^ for difference in effect size) and triglycerides (medication vs. secondary care costs P = 1.09×10^−4^) had larger effects on medication costs than secondary (or primary) care costs. Such effects reflected relative rather than absolute increases. For example, a one SD increase in SBP caused a large relative difference in annual medication costs than secondary care costs (34.18% increase (95% CI: [27.16, 41.59]) vs 8.17% increase (95% CI: [3.10, 13.49], respectively). However, the estimated absolute euro changes were similar (i.e., medication costs of €69.04 vs. secondary care costs of €69.61).

**Figure 4.**
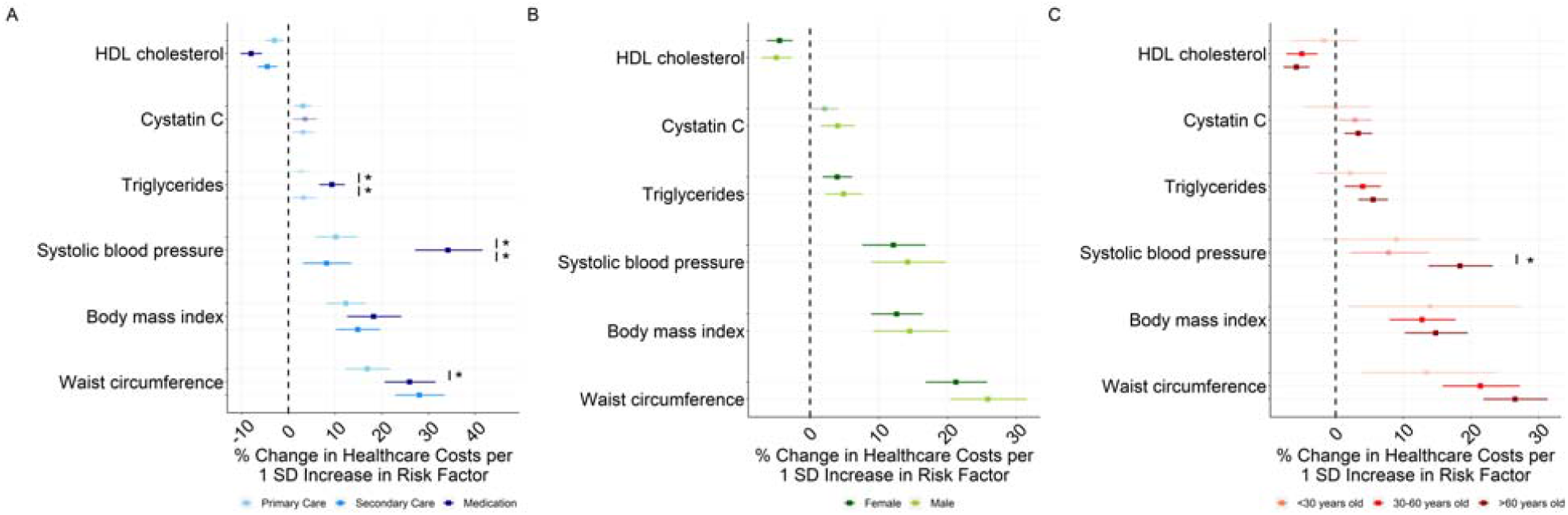
Mendelian Randomization on six biological risk factors for 373,160 FinnGen participants using the two-sample, inverse variance weighted approach stratified by (A) service type, (B) sex, and (C) age. Bars indicate 95% confidence interval. The * sign indicates significant differences between different levels of the stratification variable within the risk factor at the Bonferroni-corrected significance level (P < 8.33×10^−3^). SD is standard deviation.

We found little evidence that the relative impact of the risk factors on healthcare costs differ between females and males. Similarly, we found few differences between individuals younger than 30 years old, between 30 and 60 years old, and older than 60 years old. The only exception was a modest difference in the relative impact of SBP on healthcare costs between individuals aged 30 to 60 years old (7.79%, 95% CI: [2.12, 13.77]) compared to individuals older than 60 years old (18.38%, 95% CI: [13.71, 23.23]) for SBP (P = 3.20×10^−3^).

### Factors mediating the impact of risk factors on total healthcare costs

For the three risk factors with the largest percent change on healthcare costs (WC, adult BMI, SBP), we used MVMR to understand how much of their impact on healthcare costs can be explained by increased risk for major diseases associated with high healthcare costs (**Supplementary Table 6**). We considered the top five noncommunicable diseases from the Global Burden of Disease study:^22^ back pain, chronic ischemic heart disease, type 2 diabetes, chronic obstructive pulmonary disease, and stroke. For SBP, we additionally studied blood pressure medications as a mediator, which was not immune from collider bias but provided context for indirect effects of SBP on healthcare costs.

After accounting for the genetic effects mediated by the five noncommunicable diseases, we found that type 2 diabetes and blood pressure medications modestly mediated the effects of adult BMI and SBP on annual total healthcare costs, respectively. Adjusting for type 2 diabetes slightly attenuated the effect of adult BMI on healthcare costs from 13.64% [95% CI: 10.26, 17.12] to 10.18% [95% CI: 4.88, 15.76]. Adjusting for blood pressure medications attenuated the effect of SBP on healthcare costs from 13.08% [95% CI: 8.84, 17.48] to 4.06% [95% CI: -2.45, 10.47]. Interestingly, even after adjusting for the top five noncommunicable diseases, WC effects on healthcare costs remained similar suggesting that WC affects healthcare costs broadly beyond the increased risk of the top five major diseases.

### Replication analysis for generalizability and robustness of healthcare costs findings

We conducted several analyses to evaluate the robustness of our findings. First, we perform similar MR analysis in UK Biobank (N=307,048) and we estimated a £96.90 (€115.32) increase per SD of WC; a £94.59 (€112.57) increase per SD of adult BMI; and a £24.36 (€28.99) increase per SD of SBP. In clinical units, we estimated a £77.40 (€92.11) increase per 10 cm of WC; a £102.82 (€122.36) increase per 5 kg/m^2 of adult BMI; and a £11.77 (€14.01) increase per 10 mmHg of SBP. Similarly, in the Netherlands Twin Register (N=16,726), we estimated a €182.52 increase per SD of WC; a €264.85 increase per SD of adult BMI; and a €10.07 increase per SD of SBP. In clinical units, we estimated a €129.91 increase per 10 cm of WC; a €261.44 increase per 5 kg/m^2 of adult BMI; and a €87.69 increase per 10 mmHg of SBP. Results from these other sources are therefore in the range of our estimates, despite the different healthcare systems, data sources (e.g., different cost categories captured), and population structures (**Figure 5, Supplementary Table 3**).

**Figure 5.**
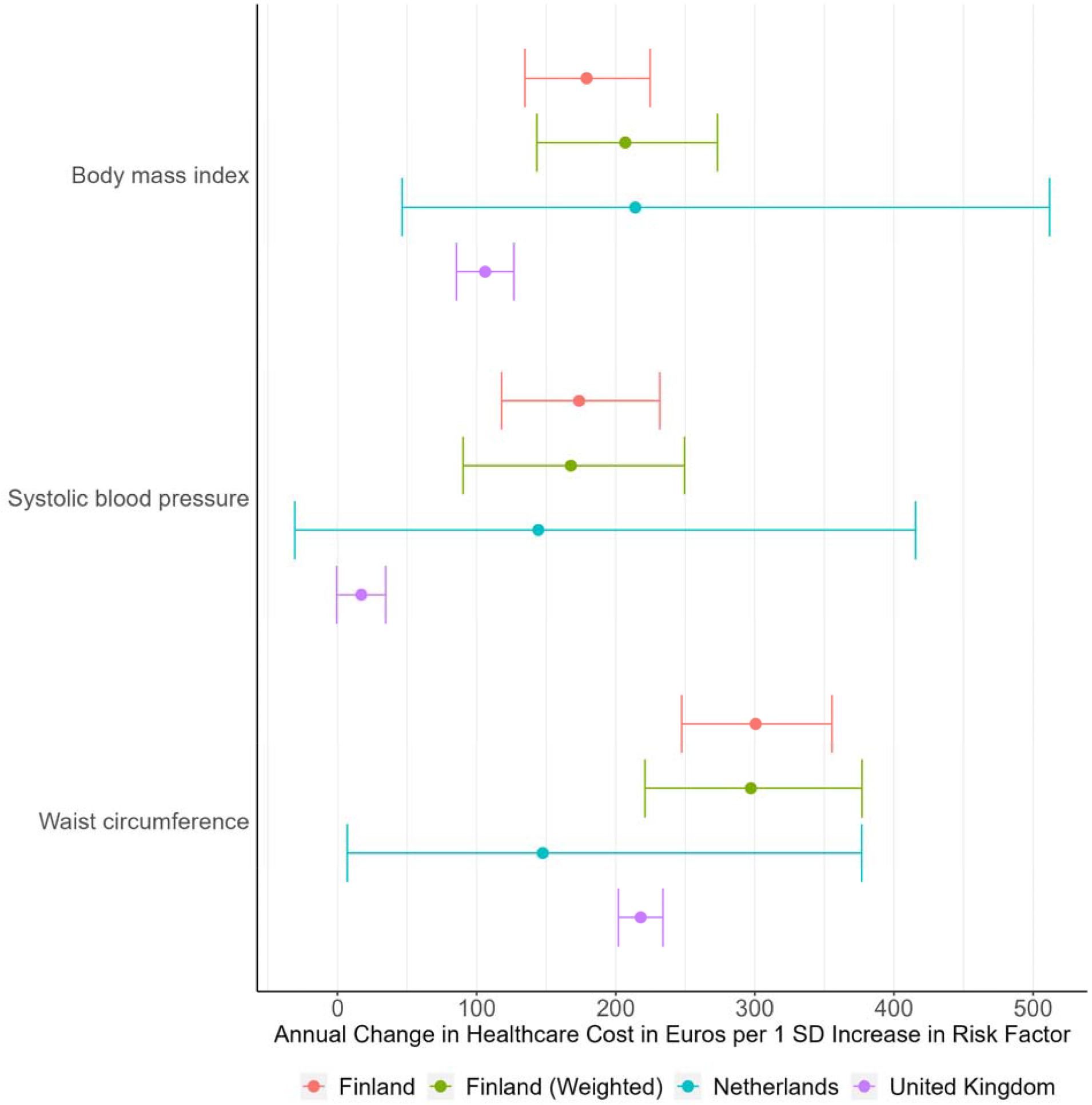
Mendelian Randomization results for total healthcare costs for 3 three main biological risk factors in a replication analysis including data from the United Kingdom (N=307,048), Netherlands (N=16,726), Finland (N=373,160) and re-weighting the FinnGen cohort to reflect the entire Finnish population. SD is standard deviation.

Second, we compared the genetic association with annual healthcare costs in FinnGen with those publicly available from the United Kingdom and Netherlands (**Supplementary Table 4**). We observed that the genetic correlation was significant between between Finland, the United Kingdom, and Netherlands. Comparing secondary care costs, Finland and the United Kingdom had a genetic correlation of 0.804 (SE = 0.05492, P = 1.61×10^−48^). Comparing primary care costs, Finland and the Netherlands had a genetic correlation of 0.7694 (SE = 0.3387, P = 2.31×10^−2^). For the Netherlands total, secondary care, and medication costs, heritability was too low to calculate genetic correlation.

Third, we calculated polygenic scores (PGS) for healthcare costs using weights from the United Kingdom (UK) and Netherlands (NL). In general, there was a large and significant association between Finnish healthcare costs and UK- and NL-based PGS, suggesting that cross-country analyses of healthcare costs may be valuable (**Supplementary Figure 1**). A 1 SD increase in the UK-based PGS for secondary care costs was associated with an increase in €128 per year (95% CI: [97, 160], P = 2.09×10^−15^) or 9.29% per year (95% CI: [8.74, 9.84], P = 6.70×10^−293^). A 1 SD increase in the NL-based PGS for total healthcare costs was associated with an increase of €15 per year (95% CI: [-19, 50], P = 3.83×10^−1^) or 2.84% per year (95% CI: [2.46, 3.21], P = 1.66×10^−50^). The lower increase observed for NL-based PGS is expected given the PGS was derived on a smaller sample size.

Finally, FinnGen is not fully representative of the general Finnish population and enriched with individuals that have been in contact with the healthcare system due recruitment being predominantly based in hospital-based settings. To evaluate the generalizability of our results to the entire Finnish population, we used inverse probability weighting with weights calculated by comparing five health and sociodemographic characteristics (i.e., age, gender, education, occupation, and region of birth) between FinnGen participants and the full Finnish population (**Supplementary Figure 2, Supplementary Table 5**). We found a high correlation between the effect sizes from the GWAS of healthcare costs and the weighted linear regression (R^2^ = 0.76). We also found similar results for the MR analysis, in which one SD increase in WC increased healthcare costs by 22.64% (95% CI: [16.84, 28.72], P = 1.53×10^−16^), one SD increase in adult BMI increased healthcare costs by 12.42% (95% CI: [6.96, 18.16], P = 4.06×10^−6^), and one SD increase in SBP increased healthcare costs by 12.56% (95% CI: [6.66, 18.80], P = 1.67×10^−5^).

## Discussion

We linked genetic information to detailed healthcare costs covering primary, secondary, and medication costs for 373,160 participants in FinnGen followed-up to a maximum of 22 years. This allowed us to evaluate the association between the genetic underpinnings of 15 clinically relevant risk factors and annual total healthcare costs. Generally, making causal inferences about the effects of these risk factors is challenging because of confounding, reverse causation, and the unfeasibility of randomized controlled trials. We address these limitations using a genetically-informed causal inference design. Under the assumptions of Mendelian Randomization, we estimated the causal effects of these risk factors on total healthcare costs. Our approach was conservative, and we chose risk factors that have strong genetic bases and high heritability. However, we did not consider important modifiable risk factors such as smoking and alcohol consumption because using MR with such risk factors represents additional challenges.

The risk factor with the largest quantitative impacts on healthcare costs were WC, followed by adult BMI and SBP. An increase of 10 cm in WC results in 15.40% increase in annual healthcare costs, which, in Finland, corresponds to approximately €202.13. The effect of WC, unlikely BMI, was not attenuated when considering the potential mediating effect of five major diseases. Previous studies have suggested that WC may be more informative than adult BMI for certain health outcomes, as WC may better reflect the accumulation of intra-abdominal fat compared with BMI.^28^ The MR study of Hazewinkel et al. in the UK Biobank found that an adverse fat distribution rather than the level of BMI may drive the relationship between BMI and higher rates of hospital admission.^29^

Importantly, we found that the impact several biomarkers (e.g., alanine aminotransferase, glycated hemoglobin, C-reactive protein, LDL cholesterol, lipoprotein(a), creatinine, vitamin D, albumin, and glucose) have on healthcare costs was modest and not significant at the Bonferroni-corrected significance level. It has been argued the MR is more valuable to reject causal claims when the genetic instrument is sufficiently strong,^30^ as in our case.

There may be two main reasons why we did not find significant effects for these biomarkers. First, elevated biomarkers can be consequences of underlying disease processes, for example, by reflecting inflammation, as in the case of C-reactive protein. Moreover, their levels can simply capture (un)healthy behaviors. For example, numerous trials have shown no benefits for Vitamin-D supplements on reducing risk for several diseases, such as cardiovascular diseases, despite supporting evidence from observational studies,^31 32^ but not from MR-based studies.^33 34 35^ Second, the effect of risk factors on healthcare costs reflects current clinical practice. If a risk factor is routinely measured and those with high levels of the risk factor are correctly targeted by preventive interventions, the increased healthcare costs associated with the preventive interventions should be counterbalanced by the reduced healthcare costs associated with the prevented disease burden. Such is the case of LDL cholesterol - if LDL cholesterol was sufficiently treated in the population, LDL cholesterol would have less of an impact on healthcare costs. Indeed, Harrison et al. observed a null impact of total serum cholesterol on other social and economic outcomes in the UK Biobank.^36^ Similarly, while glycated hemoglobin is a known marker for type 1 and 2 diabetes, proper management may results in a lower impact of glycated hemoglobin on healthcare costs as compared to a theoretical scenario where patients with high glycated hemoglobin were untreated.

Our study has limitations. First, the power and precision of our MR analysis was limited by the availability of SNPs associated with the risk factors. Second, MR is an instrumental variable analysis that uses linear model estimates that may not accurately capture non-linear effects of risk factors on healthcare costs. Third, MR uses the genetic variation assigned at conception (e.g., genetically determined risk factors), and therefore estimates the lifetime effects of risk factors on healthcare costs, rather than acute or temporary effects. For example, an intervention that reduces WC in older ages may not result in reductions in healthcare costs consistent with our estimates.

Fourth, healthcare systems worldwide vary. Finland, which has a public healthcare system, is ideal for the analysis of healthcare costs, as healthcare services are uniformly priced, and is similar to other European countries with public healthcare systems. On the other hand, countries that rely more heavily on private healthcare and insurance, such as the United States, may offer healthcare services at different costs depending on insurance plans and other factors, making the analysis of healthcare costs difficult. Moreover, our results are based on individuals of European ancestry, and genetic effects might vary across ancestry groups.

Our approach opens different research venues. Drug makers can quantify healthcare costs associated with specific drug targets, including proteins and metabolites, for which large-scale GWAS have been performed and use these results to inform drug development. Public health specialists can extend these approaches to evaluate whether screening procedures for certain biomarkers are cost-effective. Future large-scale genetic studies will likely identify genetic variants associated with healthcare costs and inform the implementation of genomic medicine approaches.

In conclusion, our results not only indicate that elevated WC, BMI and SBP are major causal contributors to healthcare costs, but could also quantify their impact on healthcare costs within a causal inference framework. This has implications for the cost-effectiveness of interventions and policies that influence these biomarkers. Several other biomarkers routinely measured in clinical setting are unlikely to directly impact on healthcare costs, either because they are not causal to healthcare cost, or because they are already well managed in the clinic.

## Data Availability

All data produced in the present study are available upon request to the authors.

## Acknowledgments

We would like to acknowledge Camiel M. van der Laan and Dorret I. Boomsma for their contribution of healthcare costs summary statistics from the Netherlands Twin Register, as well as Bart Ferket for his insights on health economics in designing this study. We would also like to acknowledge all of the study participants for their generous participation in FinnGen and other biobanks, as well as FinnGen as a study group that has contributed to this study.

## Supplementary Materials

*Supplementary Table 1. Summary of summary statistics used as genetic instruments for exposures and mediators*.

*Supplementary Table 2. Full Mendelian Randomization results for all risk factors using all methods in Finland*.

*Supplementary Table 3. Mendelian Randomization replication results in the United Kingdom and Netherlands*.

*Supplementary Table 4. Genetic correlation between Finland, United Kingdom, and Netherlands*.

*Supplementary Table 5. Mendelian Randomization results for reweighted FinnGen cohort*.

*Supplementary Table 6. Multivariable Mendelian Randomization results*.

**Supplementary Figure 1.**
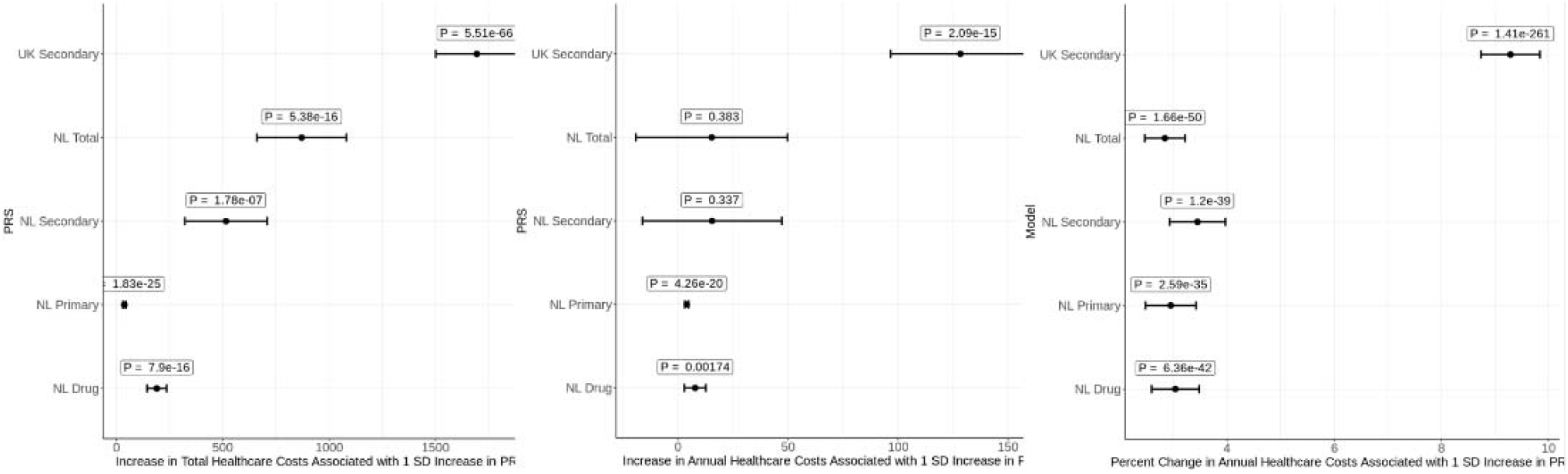
Polygenic score analysis between Finland, United Kingdom, and Netherlands.

**Supplementary Figure 2.**
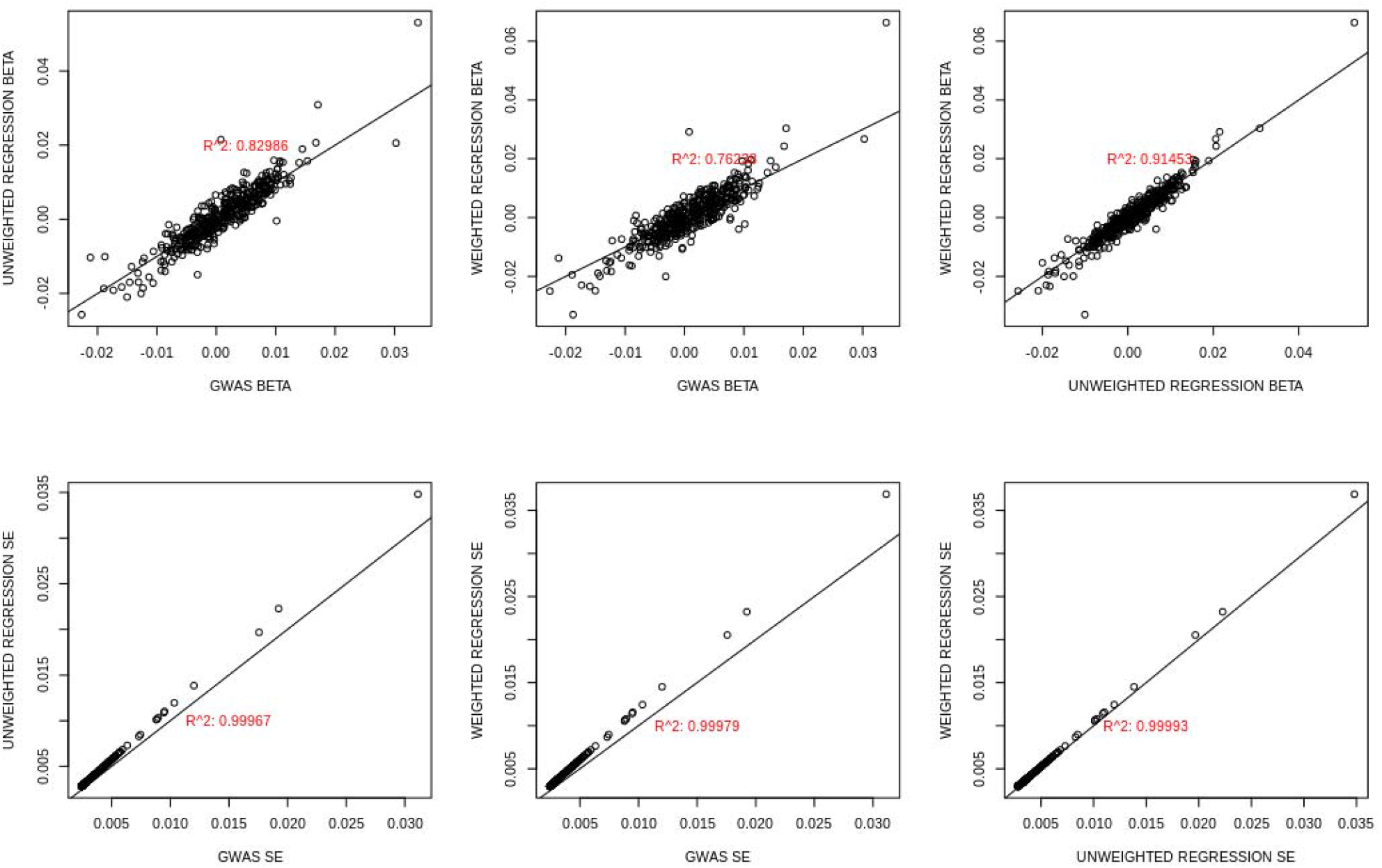
Correlation between GWAS- and weighted linear regression-based models in reweighted FinnGen cohort.

## References

1. Organization WH. Global spending on health: Weathering the storm. In:2020.

2. Edemekong PF, Tenny S. Public Health. In:2021.

3. Gallet CA, Doucouliagos H. The impact of healthcare spending on health outcomes: A meta-regression analysis. Soc Sci Med. 2017;179:9–17.

4. Bolnick HJ, Bui AL, Bulchis A, et al. Health-care spending attributable to modifiable risk factors in the USA: an economic attribution analysis. Lancet Public Health. 2020;5(10):e525–e535.

5. Goetzel RZ, Henke RM, Head MA, Benevent R, Rhee K. Ten Modifiable Health Risk Factors and Employees’ Medical Costs-An Update. Am J Health Promot. 2020;34(5):490–499.

6. Meraya AM, Raval AD, Sambamoorthi U. Chronic condition combinations and health care expenditures and out-of-pocket spending burden among adults, Medical Expenditure Panel Survey, 2009 and 2011. Prev Chronic Dis. 2015;12:E12.

7. Dieleman JL, Cao J, Chapin A, et al. US Health Care Spending by Payer and Health Condition, 1996-2016. JAMA. 2020;323(9):863–884.

8. Dixon P, Sallis H, Munafo M, Davey Smith G, Howe L. The causal effect of cigarette smoking on healthcare costs. In:2022.

9. Smith GD, Ebrahim S. ‘Mendelian randomization’: can genetic epidemiology contribute to understanding environmental determinants of disease? Int J Epidemiol. 2003;32(1):1–22.

10. Dixon P, Hollingworth W, Harrison S, Davies NM, Davey Smith G. Mendelian Randomization analysis of the causal effect of adiposity on hospital costs. J Health Econ. 2020;70:102300.

11. Kurz CF, Laxy M. Application of Mendelian Randomization to Investigate the Association of Body Mass Index with Health Care Costs. Med Decis Making. 2020;40(2):156–169.

12. Harrison S, Dixon P, Jones HE, Davies AR, Howe LD, Davies NM. Long-term cost-effectiveness of interventions for obesity: A mendelian randomisation study. PLoS Med. 2021;18(8):e1003725.

13. Dixon P, Harrison S, Hollingworth W, Davies NM, Davey Smith G. Estimating the causal effect of liability to disease on healthcare costs using Mendelian Randomization. Econ Hum Biol. 2022;46:101154.

14. Kurki MI. FinnGen: Unique genetic insights from combining isolated population and national health register data. In:2022.

15. Mäklin S, Bonfire P. Unit costs of health and social care in Finland in 2017. In:2021.

16. Kela. Finnish statistics on medicines. In:2020.

17. Hemani G, Zheng J, Elsworth B, et al. The MR-Base platform supports systematic causal inference across the human phenome. Elife. 2018;7.

18. Mbatchou J, Barnard L, Backman J, et al. Computationally efficient whole-genome regression for quantitative and binary traits. Nat Genet. 2021;53(7):1097–1103.

19. Hartwig FP, Davies NM, Hemani G, Davey Smith G. Two-sample Mendelian randomization: avoiding the downsides of a powerful, widely applicable but potentially fallible technique. Int J Epidemiol. 2016;45(6):1717–1726.

20. Hemani G, Tilling K, Davey Smith G. Orienting the causal relationship between imprecisely measured traits using GWAS summary data. PLoS Genet. 2017;13(11):e1007081.

21. Burgess S, Thompson SG. Multivariable Mendelian randomization: the use of pleiotropic genetic variants to estimate causal effects. Am J Epidemiol. 2015;181(4):251–260.

22. Collaborators GDaI. Global burden of 369 diseases and injuries in 204 countries and territories, 1990-2019: a systematic analysis for the Global Burden of Disease Study 2019. Lancet. 2020;396(10258):1204–1222.

23. Dixon P, Davey Smith G, Hollingworth W. The Association Between Adiposity and Inpatient Hospital Costs in the UK Biobank Cohort. Appl Health Econ Health Policy. 2019;17(3):359–370.

24. de Zeeuw EL, Voort L, Schoonhoven R, et al. Safe Linkage of Cohort and Population-Based Register Data in a Genomewide Association Study on Health Care Expenditure. Twin Res Hum Genet. 2021;24(2):103–109.

25. Bulik-Sullivan B, Finucane HK, Anttila V, et al. An atlas of genetic correlations across human diseases and traits. Nat Genet. 2015;47(11):1236–1241.

26. Ge T, Chen CY, Ni Y, Feng YA, Smoller JW. Polygenic prediction via Bayesian regression and continuous shrinkage priors. Nat Commun. 2019;10(1):1776.

27. Chang CC, Chow CC, Tellier LC, Vattikuti S, Purcell SM, Lee JJ. Second-generation PLINK: rising to the challenge of larger and richer datasets. Gigascience. 2015;4:7.

28. Ross R, Neeland IJ, Yamashita S, et al. Waist circumference as a vital sign in clinical practice: a Consensus Statement from the IAS and ICCR Working Group on Visceral Obesity. Nat Rev Endocrinol. 2020;16(3):177–189.

29. Hazewinkel AD, Richmond RC, Wade KH, Dixon P. Mendelian randomization analysis of the causal impact of body mass index and waist-hip ratio on rates of hospital admission. Econ Hum Biol. 2022;44:101088.

30. Davey Smith G, Hemani G. Mendelian randomization: genetic anchors for causal inference in epidemiological studies. Hum Mol Genet. 2014;23(R1):R89–98.

31. Dobnig H, Pilz S, Scharnagl H, et al. Independent association of low serum 25-hydroxyvitamin d and 1,25-dihydroxyvitamin d levels with all-cause and cardiovascular mortality. Arch Intern Med. 2008;168(12):1340–1349.

32. Judd SE, Tangpricha V. Vitamin D deficiency and risk for cardiovascular disease. Am J Med Sci. 2009;338(1):40–44.

33. Brøndum-Jacobsen P, Benn M, Afzal S, Nordestgaard BG. No evidence that genetically reduced 25-hydroxyvitamin D is associated with increased risk of ischaemic heart disease or myocardial infarction: a Mendelian randomization study. Int J Epidemiol. 2015;44(2):651–661.

34. Manousaki D, Mokry LE, Ross S, Goltzman D, Richards JB. Mendelian Randomization Studies Do Not Support a Role for Vitamin D in Coronary Artery Disease. Circ Cardiovasc Genet. 2016;9(4):349–356.

35. Jiang X, Ge T, Chen CY. The causal role of circulating vitamin D concentrations in human complex traits and diseases: a large-scale Mendelian randomization study. Sci Rep. 2021;11(1):184.

36. Harrison S, Davies AR, Dickson M, et al. The causal effects of health conditions and risk factors on social and socioeconomic outcomes: Mendelian randomization in UK Biobank. Int J Epidemiol. 2020;49(5):1661–1681.

